# Mitigation policies and vaccination in the COVID-19 pandemic: a modelling study

**DOI:** 10.1101/2021.01.27.21250651

**Authors:** Naiara C.M. Valiati, Daniel A.M. Villela

## Abstract

The perspective of vaccination to protect human population from infection of SARS-CoV-2 virus has great potential to control the pandemic. Nevertheless, vaccine planning requires phased introduction with age groups, health workers, and vulnerable people. We developed a mathematical model capable of capturing the dynamics of the SARS-CoV-2 dissemination aligned with social distancing, isolation measures, and vaccination. The city of Rio de Janeiro provides a case study to analyze possible scenarios including non–pharmaceutical interventions and vaccination in the epidemic scenario. Our results shows that a combination of different policies such as case isolation and social distancing are more effective for mitigating the epidemics. Furthermore, these policies will still be necessary in a phased vaccination program. Therefore, health surveillance activities should be maintained along with vaccination planning in scheduled groups until a large vaccinated coverage is reached.

## 1 Introduction

Since the emergence of the SARS-CoV-2 virus, the COVID-19 pandemic reached many countries causing millions of severe cases and deaths. The need of interventions were necessary to mitigate the pandemic by reducing dissemination and in the next phase by starting vaccination. Even with a phased vaccination, some measures remain important such as social distancing and isolation of cases until large vaccine coverage is achieved.

A number of models studied the impact of social distancing [1]. Models range from understanding the epidemiological mechanisms behind SARS-CoV-2 and also to predict the dynamics of epidemic. A schematic review by Wynants evaluates diverse models against their predictive capabilities [2].

Among the effects expected from these interventions, the first one is delaying the peak of the pandemic, which is important in order to healthcare organizations better prepare methods, personnel and equipment to deal with the coming hospitalizations and cases. The second and most wanted is flattening the pandemic curve once it starts, the goal of this objective is to open ease the resources allocation and to give more time to the healthcare system to handle the pandemic and its effects. Vaccination should provide the approach for controlling it, depending on the vaccine efficacy.

In this work, we evaluate how different scenarios of interventions compare to each other in order to better deal with the ongoing SARS-CoV-2 pandemic. Results also consider the city of Rio de Janeiro, Brazil, as case study. Implications of the results, however, are general, such that could be extended to other similar cities. Also, as non-pharmaceutical measures are key to mitigate effects of the pandemic, the perspective of controlling it comes with vaccination. Yet, its policies and methods for application need yet to be systematically addressed. At present, there is already the surge of a second-wave in many countries [3, 4]. Health surveillance should be maintained along with the planning for effective vaccination.

## 2 Methods

### 2.1 Model

We modeled different scenarios with an ODE-based compartmental model. In the model, susceptible individuals (S) can evolve to exposed (E) conditions when in contact with infected individuals. The group of infected individuals is divided between asymptomatic cases (Y), symptomatic cases (C), which includes both mild and moderate cases, and severe cases (H). This last group occurs from the evolution of the symptomatic group and, therefore, is considered hospitalized. All infected individuals can evolve to death (D) or recovered (R). Each one of these model classes is stratified by age group ranging from 5 to 5 years, from 0 to 100 years, added by one last age group of higher than 100 years, in a total of 21 groups of distinct age ranges for each compartment of the model.

We also included the possibility of vaccination in our model. Susceptible individuals are vaccinated at a coverage rate of *η*. These vaccinated individuals will take a pre-determined time *τ*_*immun*_ to develop immunity at a probability of *ρ*_*I*_ when they evolve to immunized status (I). Due to incomplete vaccine efficacy, we included the possibility of the vaccinated individual not developing the required immunization and, therefore, being a susceptible-but-deemed-immunized individual (Im). These individuals are in a separate compartment apart from susceptible individuals because the former might lessen their social distancing and isolation measures, thinking that now they are immunized and somewhat away from the danger.

The infection rate between susceptible individuals and symptomatic is *β*, and with asymptomatic individuals is *β*_*A*_. When they become exposed individuals, the time to evolve to infected is the incubation time *τ*_*inc*_. At the end of this time, the individual has a probability *ρ*_*S*_ of developing symptoms.

The time for an asymptomatic individual might evolve to death is 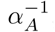, whereas for the symptomatic cases is *α* ^*−*1^, *α* > *α*_*A*_ due to expected morbidity in these groups, including that asymptomatic individuals do not present themselves as clinical cases. The symptomatic individual can evolve to a severe case with a risk probability of *α*_*H*_.

The recovery of infected individuals (symptomatic and severe) is controlled by the recovery rate *γ*, being modified to *γ*_*H*_ in the case of severe cases. Severe case are hospitalized and thus receiving proper assistance confronting the sickness. The hospitalized individual can recover after a determined period, controlled by the discharge time *τ*_*disc*_ and dyspnea time *τ*_*disp*_.

The ODE system which resumes this model is:

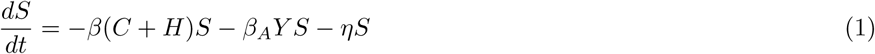

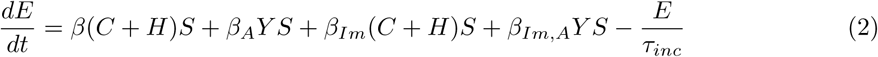

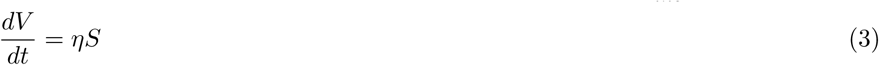

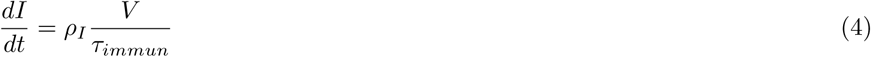

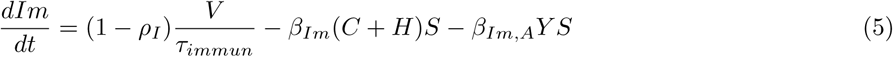

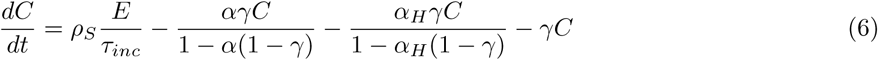

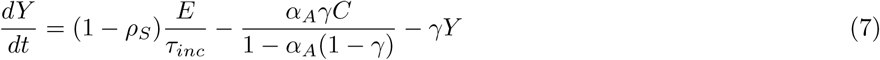

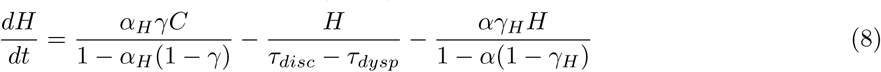

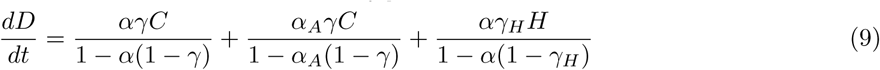

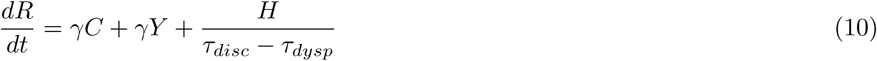

#### 2.1.1 Social distancing interventions

The model enables the application of intervention measures with the social distancing of specific age groups. Social distancing affects people in reducing the probability of encounters between infected and susceptible individuals. Thus, we simulate this condition by reducing the infection rates *β, β*_*A*_, *β*_*I*_ and *β*_*Im*_ for the specific age groups. Due to imperfect application of social distancing intervention, each intervention is controlled by a success rate.

The fact that the model is stratified by age groups opens a new range of different scenarios, e.g., when applying the social distancing intervention to younger age groups, we can simulate a closed schools condition. The reduction is applied to the *R*_0_, from which the infection rates are calculated, value multiplying it by the reduction factor *κ* with a pre-defined value. The social distancing applied to the 0-20 years old age groups is labeled SD-Y, when applied to the age groups higher than 60 years old is labeled SD-E, and when we apply the reduction to all age groups, we label this condition as SD-A.

#### 2.1.2 Isolation interventions

The application of isolation interventions is made by reducing the encounter probability between susceptible and infected individuals. Different scenarios are tested in this work. In the lockdown scenario (L), we alter the susceptible flow equation to

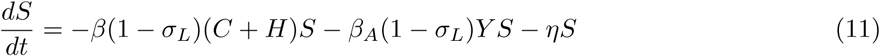

Another intervention possibility is when tests are applied to the individuals, and a quarantine is applied where symptomatic cases are isolated with a probability *σ* and asymptomatic with a probability *σ*_*A*_, this condition is labeled as TQ-C. In this scenario, we modify the susceptible flow equation to

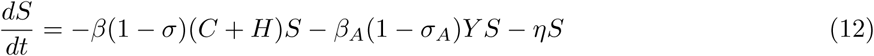

If we only isolate the severe cases (scenario TQ), we change the susceptible individuals flow equation to

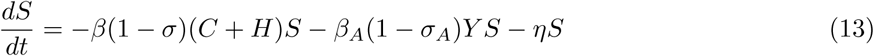

The scenario where we only isolate the severe cases is termed TS, and we modify the susceptible flow equation to

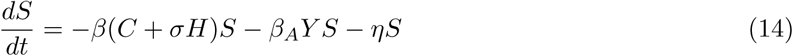

The exposed, vaccinated, and falsely immunized are changed just like the susceptible flow, depending on the scenario.

The Table 1 summarizes the parameters used in the model with their respective values and references.

**Table 1:** Description of parameters in the model and values used in simulations with references, if available.

| Parameter | Description | Value |
| --- | --- | --- |
| $\beta$ | Infection rate | Calculated using $R_0$ |
| $f_A$ | Asymptomatic factor | 0.42 [5] |
| $\beta_A$ | Asymptomatic infection rate | $f_A \cdot \beta$ |
| $\sigma$ | Probability of successful isolation of symptomatic individuals | 0.30 – 0.80 |
| $\sigma_A$ | Probability of successful isolation of asymptomatic individuals | 0.20 |
| $\sigma_L$ | Probability of successful isolation during lockdown | 0.75 |
| $\rho_S$ | Probability of developing symptoms | 0.83 [5] |
| $\alpha$ | Death risk | Depends on age group [6] |
| $\alpha_H$ | Hospitalization risk | Depends on age group [7] |
| $\tau_{dysp}$ | Time for dyspnea | 7 days [8] |
| $\tau_{disc}$ | Discharge time | 22 days [8] |
| $\tau_{inc}$ | Incubation time | 5.1 days [9] |
| $\gamma$ | Recovery rate | 1/6.5 |
| $\gamma_H$ | Recovery rate for hospitalized individuals | Calculated using $\tau_{disc}$ and $\tau_{dysp}$ |

The parameter *β* is calculated from the previous definition of *R*_0_ value, the asymptomatic value *f*_*A*_, the probability of developing symptoms *ρ*_*S*_, and the incubation time *τ*_*inc*_ with

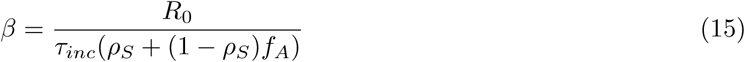

The infection rate regarding asymptomatic individuals is obtained by the product of *β* with *f*_*A*_, while the infection rate regarding asymptomatic individuals is obtained from the product of *β* with *f*_*I*_. Fig. 1 depicts a schematic diagram showing the model compartments.

**Figure 1:**
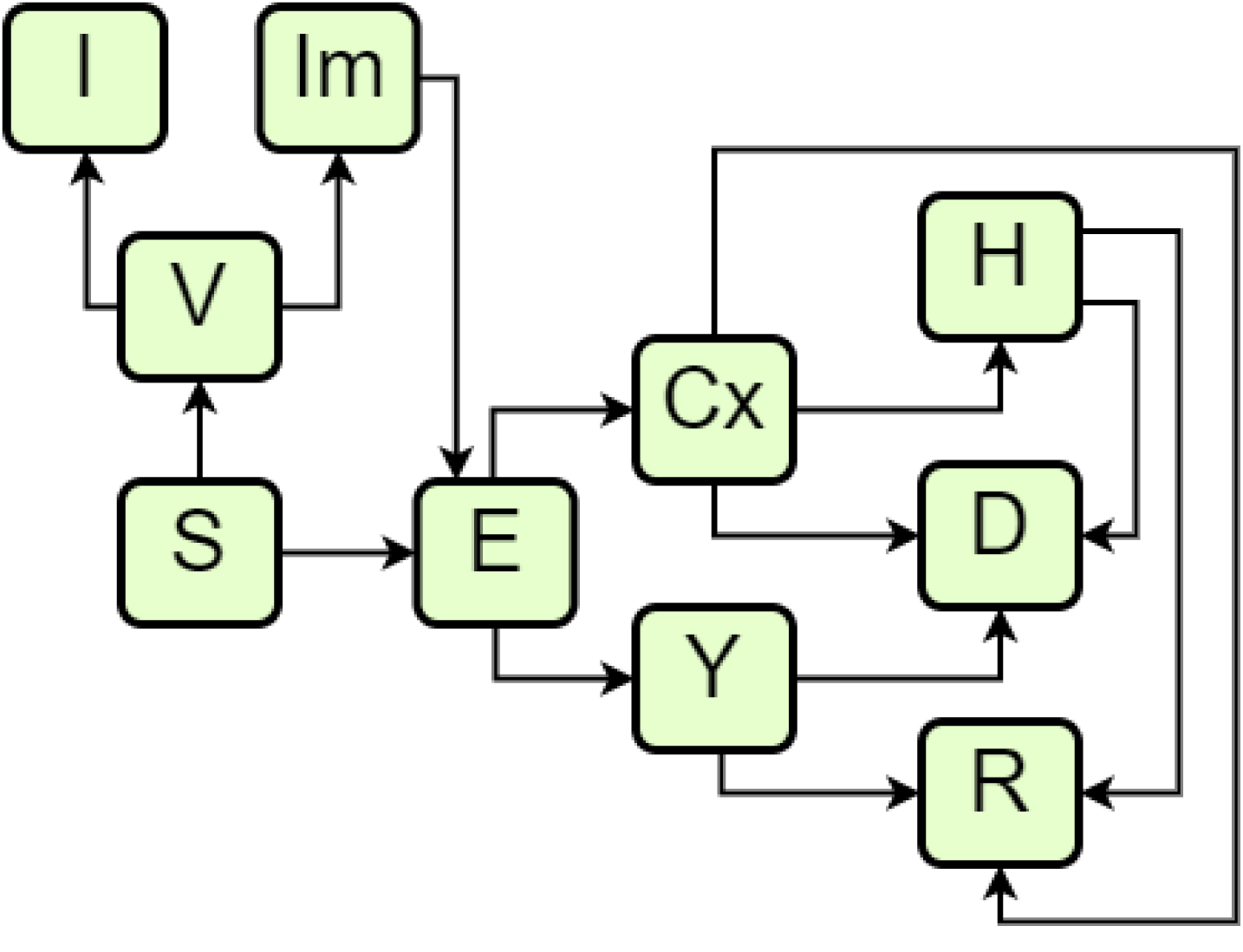
Schematic diagram of the model compartments.

#### 2.1.3 Stochastic implementation

The model is implemented with a discrete-time fashion. Typically, for each time step all transitions are evaluated as probabilities and the number flowing from a compartment to the other linked compartments, including keeping the state, are drawn from a multinomial distribution.

### 2.2 Case study

Parameters of the model were adjusted to number of cases and the dynamics observed in the municipality of Rio de Janeiro. Data from Severe Acute Respiratory Illness (SARI) are compared to the results of new daily hospitalizations. In contrast, data from Acute Respiratory Illness (ARI) notified cases are compared to the results of new daily cases. All notified data is retrieved from the public database OpenDataSus (available at https://opendatasus.saude.gov.br/dataset).

Throughout the pandemic, the scenario was altered several times due to governmental decisions of applying the interventions or making them more flexible and the incomplete adherence of the population. In this section, we evaluate how the model behaves when we use the same quarantine severity as applied by the government for each period while comparing the results to real-time data. Our approach is based on the Rio de Janeiro municipality and state real pandemic decrees, with small adjustments. We consider no intervention done between 01 January 2020 and 15 March 2020 (day 1 to day 74). Starting from 16 March 2020 until 27 March 2020 (day 75 to day 86), we consider that this is the beginning of the pandemic, where the government started to apply some intervention measures. However, the population was still not impacted by the severity of the pandemic. Therefore we considered the social distancing of young age and elderly groups, together with the quarantine of only severe cases.

From 28 March 2020 to 4 April 2020 (day 87 to day 107), social distancing becomes more widespread, and therefore we consider a social distancing of all age groups. From 5 April 2020 to 14 May 2020, the interventions become more restrictive, and now the isolation of cases extends to symptomatic cases when confirmed by testing. A higher isolation rate is considered between the dates of 15 May 2020 and 29 May 2020. On 30 May 2020, up to 2 June 2020, intervention measures start to loosen, and the isolation is loosened to the isolation of symptomatic, asymptomatic, and severe cases. To reflect this change, the probability of successful isolating symptomatic cases varies from 0.75 to 0.60. Proceeding with flexibilization, from 3 June 2020 to 12 July 2020, the quarantine of asymptomatic cases is abandoned. The last intervention change is from 13 July 2020 to 31 October 2020, when social distancing is loosened to only social distancing of young and elderly groups.

To better fit the model to the real notification data, we estimated *R*_0_ = 2.6, the reduction factor of the social distancing to be 0.72, the success in isolating symptomatic cases to be 0.60, while 0.20 for the asymptomatic cases. Also, we considered that the first cases were imported on 11 February 2020.. Reporting rate of severe cases (SARI) are 96% of the real cases, accounting for small under-reporting, whereas under-reporting of notified ARI disease cases are 20% of the actual number of ARI cases.

The number of SARI cases notified in the city of Rio de Janeiro, daily aggregated, is evaluated from January to the end of October of 2020. Regarding the ARI notified cases, the data is evaluated from January to the end of September of 2020. This data range is considered an acceptable range to avoid the effect of dramatic sub notification due to notification delay.

### 2.3 Vaccination Program

Vaccination schedules are still being studied for SARS-CoV-2, with different strategies being applied due to diverse factors [10]. However, we expect that this schedule might closely follow other respiratory syndromes’ vaccination, like influenza. In this work, we will consider an effective, tight, and compromised vaccination program to assess if this would be enough to halt the pandemic effectively. Therefore, our vaccination schedule is comprised of 4 phases, each lasting for 15 days. In the first phase, individuals older than 60 years old are vaccinated at a rate of 1.0%. Critical risk workers and people with comorbidities and underlying conditions (in the age of 21 to 59 years old) are not specifically modeled but these other adult ages are vaccinated at 0.1% per day. In the second phase, more young adults are included (more critical risk workers, school staff, individuals with disabilities), raising the group’s vaccination rate to 0.5%. In the third phase, all young adults and children are included at a rate of 1.0% per day, whilst individuals older than 60 years old are vaccinated at a rate of 0.1% per day. In the fourth and final phase, individuals of all groups are allowed to be vaccinated at a rate of 1.0% per day.

## 3 Results

In order to better evaluate the effect of different interventions separately, we compare the number of daily deaths and daily hospitalizations for different interventions using a population of the size of the city of Rio de Janeiro.

As shown in Fig.2, there is a marked difference in the effectiveness of each intervention alone. All the cases in which social distancing was applied alone had a less pronounced effect than the quarantine of cases scenarios, except for the quarantine of only the severe cases (TS), which had a minimal delaying effect at the peak. A combination of mitigation policies makes significant impact in the peak of number of cases.

**Figure 2:**
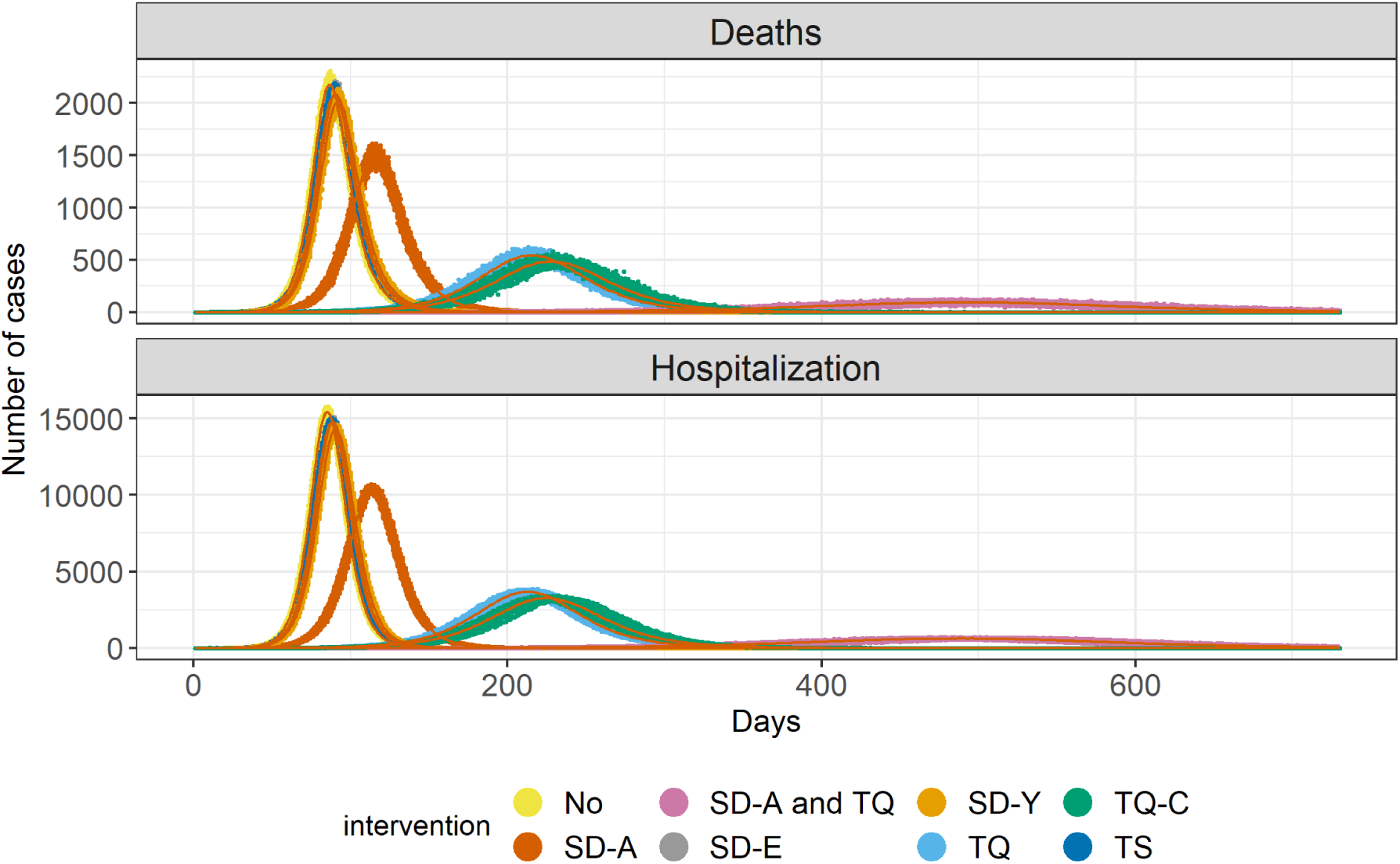
Different scenarios model comparison. A different color identifies each intervention. The points represent the stochastic calculation done with the model considering the given probabilities with 100 iterations per day. The red lines are means of each intervention. The used parameters are given in Table 1, with the exception of *R*_0_, which is 3.5.

As our model is stratified by age groups, we also observe how the different interventions change the number of deaths and hospitalizations by age, as shown by Fig.3. The quarantine of all cases, the social distancing of all individuals, and the combination of this intervention with the quarantine of symptomatic cases are the three most effective interventions, as also seen by Fig.2. In all cases, despite isolating or distancing different age groups, the pattern of hospitalizations and deaths regarding age groups is very similar. The major difference is observed in delaying the pandemic peak and the pandemic’s length, broadening its profile through time, but not through age groups. Hospitalizations are centered around older groups, mainly individuals around 60 years old and older, in all interventions. Concerning hospitalizations, there is a more even distribution among age groups. However, it is essential to note that hospitalizations of young age individuals (younger than 30 years old) are considerably pronounced.

**Figure 3:**
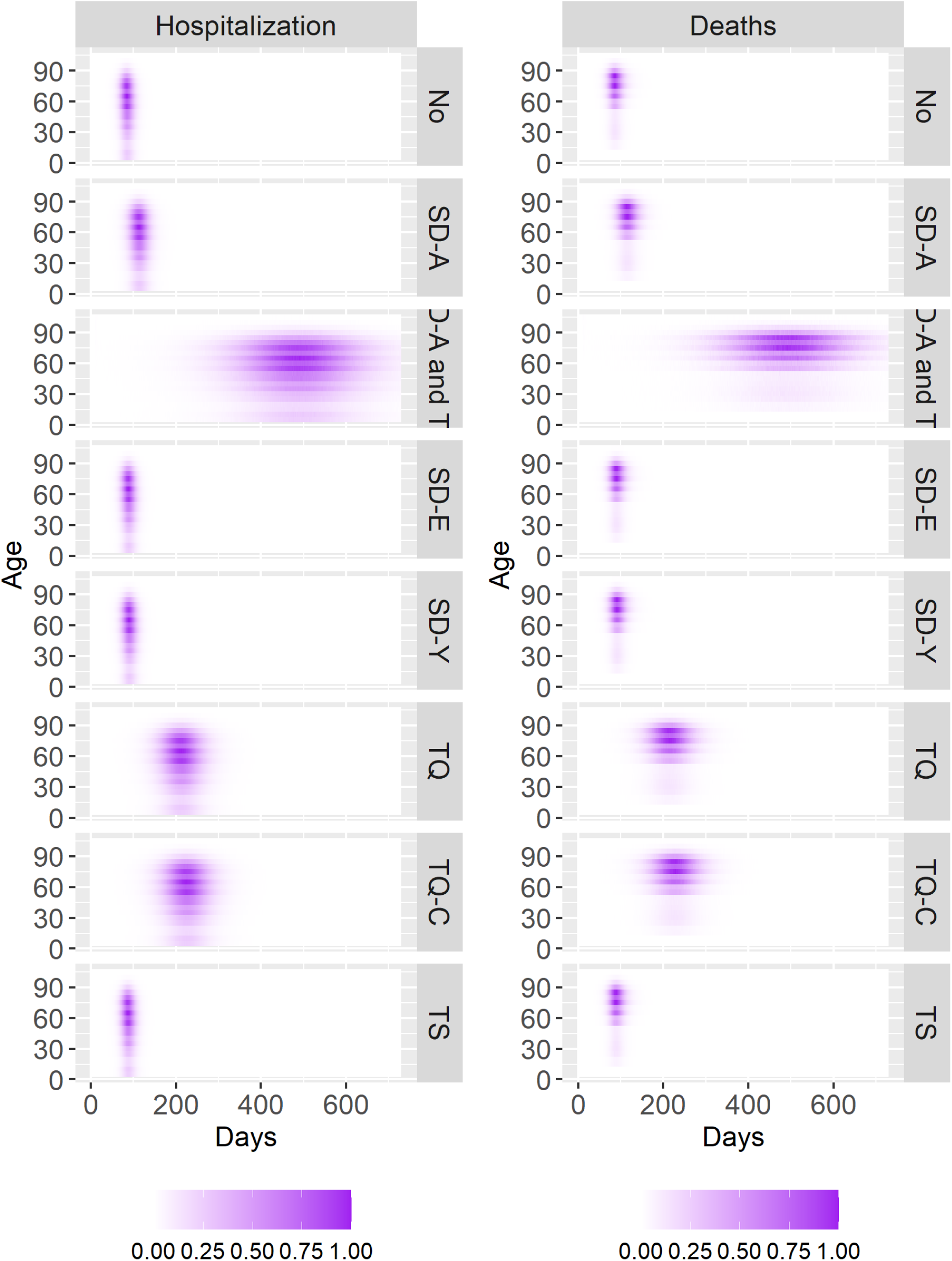
Normalized death and hospitalization profiles for different intervention scenarios. Normalized values are calculated by the quotient of each daily new hospitalization or death by the highest hospitalization or death of the group with most hospitalizations or death through the pandemic.

Also, in Figure 3, despite profile similarity across age groups, some age groups are more affected since the beginning of the pandemic and at the end. There is a distortion of the profile’s rectangular shape observed in almost all scenarios, in favor of a more oval-oriented shape, which is more pronounced in the SD-A and TQ, only TQ, and only TQ-C scenarios.

Regarding the case study of Rio de Janeiro municipality, as shown by Fig. 4, the model presents an excellent fit given number of SARI and ARI cases. The model captured the dynamics in Rio de Janeiro successfully regarding the hospitalizations compared to SARI notified cases. Regarding the influenza-like illness, the model does not account for all influenza-like illness, but it is limited to SARS-CoV-2 cases. Therefore, its reporting parameter being only 0.80.

**Figure 4:**
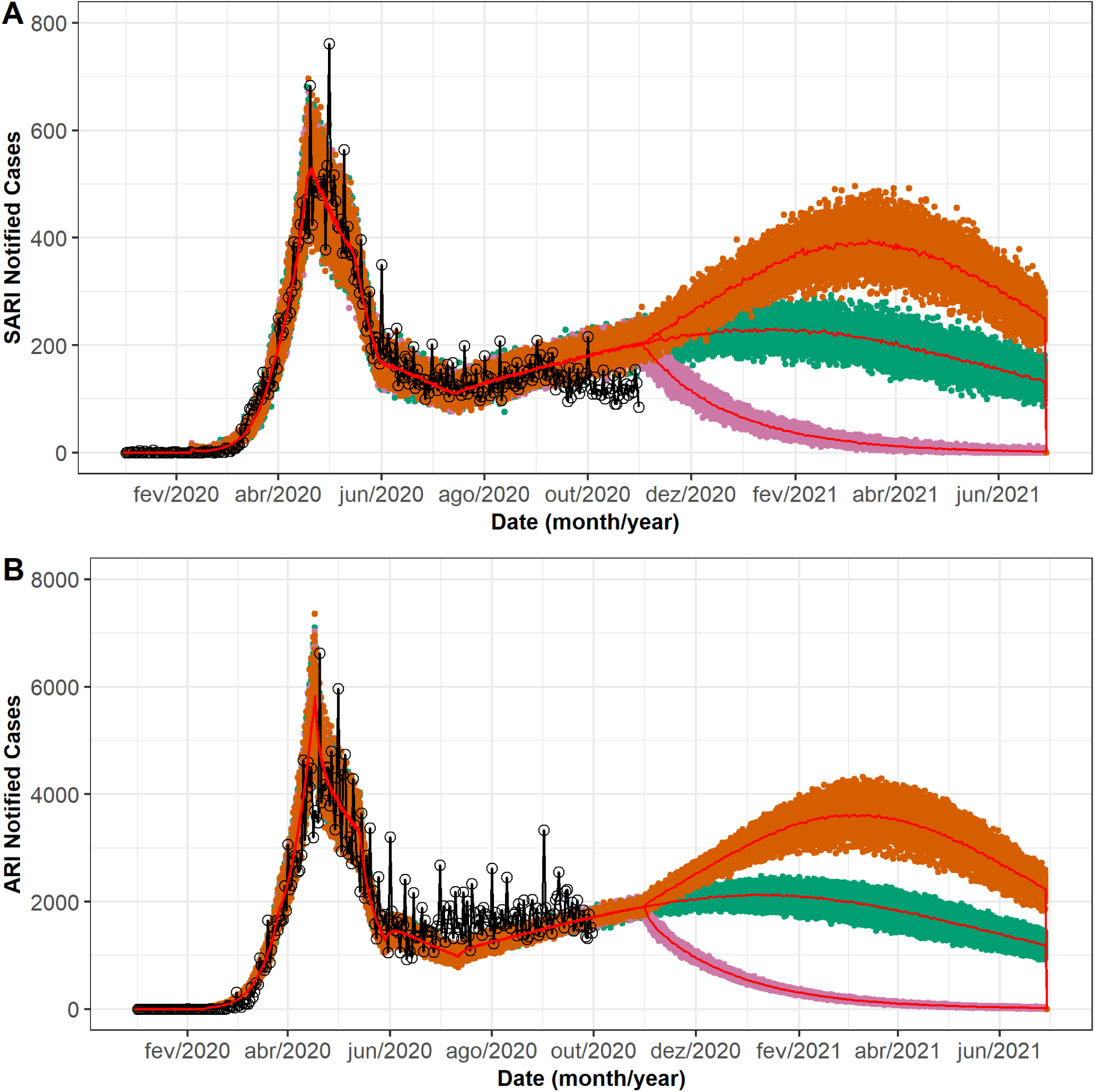
Model results for new daily hospitalizations and cases of SARI in Rio de Janeiro (A). Model results for new daily symptomatic cases and ARI notified cases in Rio de Janeiro (B). Notified cases of SARI and ARI in Rio de Janeiro are represented by black lines, other colors represent the different simulated scenarios: social distancing is abandoned (orange), the quarantine strengthening together with the social distancing of all groups and isolation of both symptomatic and asymptomatic individuals (pink), and current social distancing with symptomatic isolation scenario is maintained (green). Red lines represents the median values in each scenario.

The results from evaluating different scenarios of flexibilization are shown in Fig.4. Maintaining the actual quarantine and social distancing status as of the end of October still gives the pandemic the chance to be on the rise, reaching a peak only in few months. If the social distancing is abandoned, then there is a stronger rise in the pandemic. Halting the cases rise and changing its course happens when both the quarantine and social distancing are enforced.

Even though a second peak is not yet reached in Rio de Janeiro as of now by the official notified data, it is seen in Fig. 4 that ARI notified cases are on the rise.

Fig.5 shows the comparison between two vaccination dates to be studied. As expected, the sooner the vaccination program begins, the sooner the number of new daily hospitalizations and deaths start to fall. However, it is worth mentioning that social distancing and quarantine are still maintained in all three scenarios.

**Figure 5:**
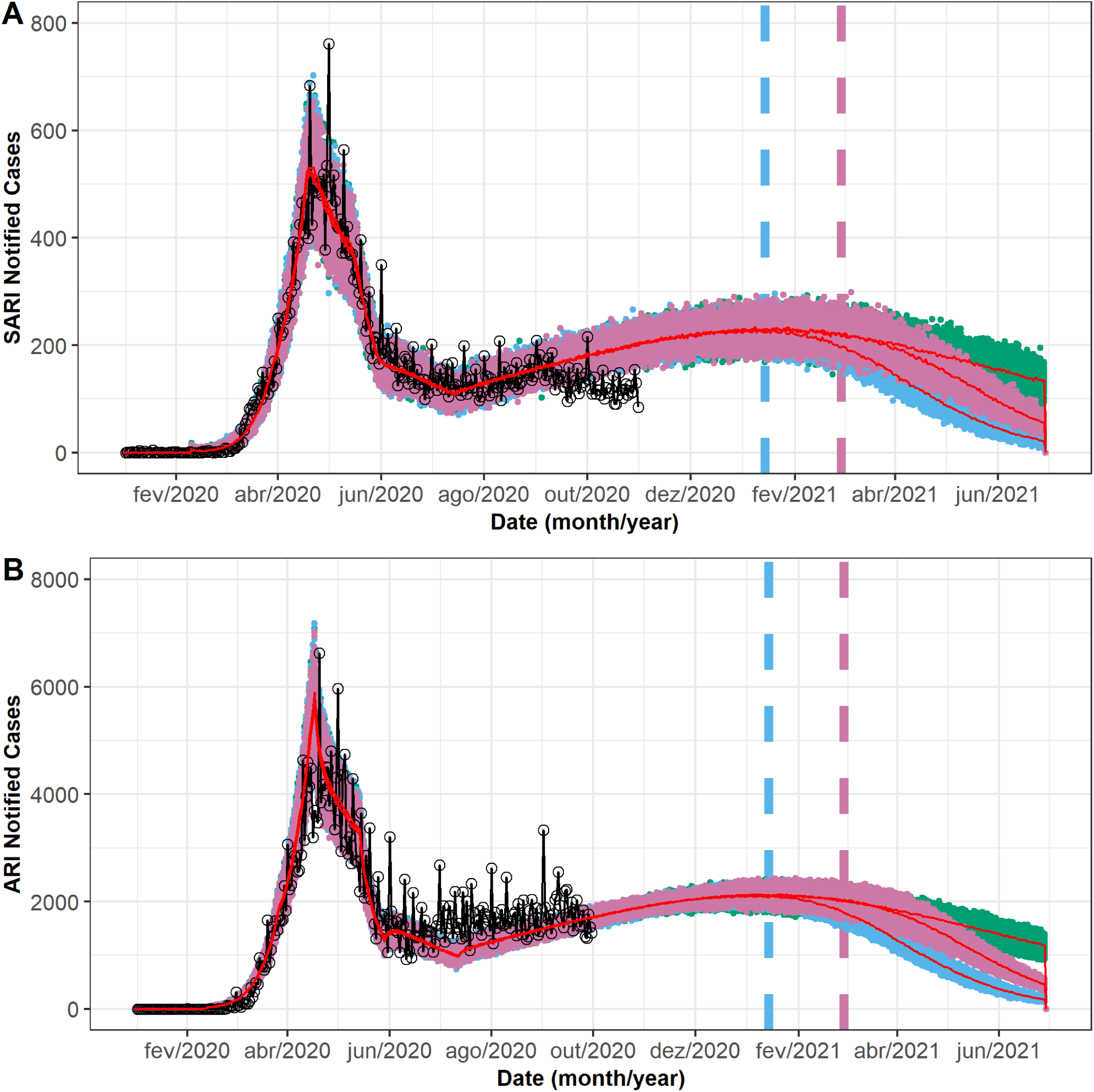
Model results and cases of SARI and ARI in Rio de Janeiro accounting for vaccination events. The black line represents notified cases of SARI and ARI in Rio de Janeiro. The other symbols represent the different simulated scenarios calculated by the model. The blue symbols represent a scenario in which vaccination begins on 15 January 2020. The pink symbols represent a scenario in which vaccination begins on 1 March 2020, while the green symbols represent a scenario without vaccination

To answer the question of how the pandemic would behave if we modified the intervention status during the vaccination rollout, we simulated the conditions presented at Fig.6. Fig.6 also shows that in the scenarios in which social distancing and quarantine are present, and in the case of only the quarantine is considered, vaccination plays a fundamental role. When vaccination starts, this downfall is advanced and accelerated, which is evidenced by the observed inflection point. Abandoning social distancing, however, generates an increase in the number of expected SARI cases.

**Figure 6:**
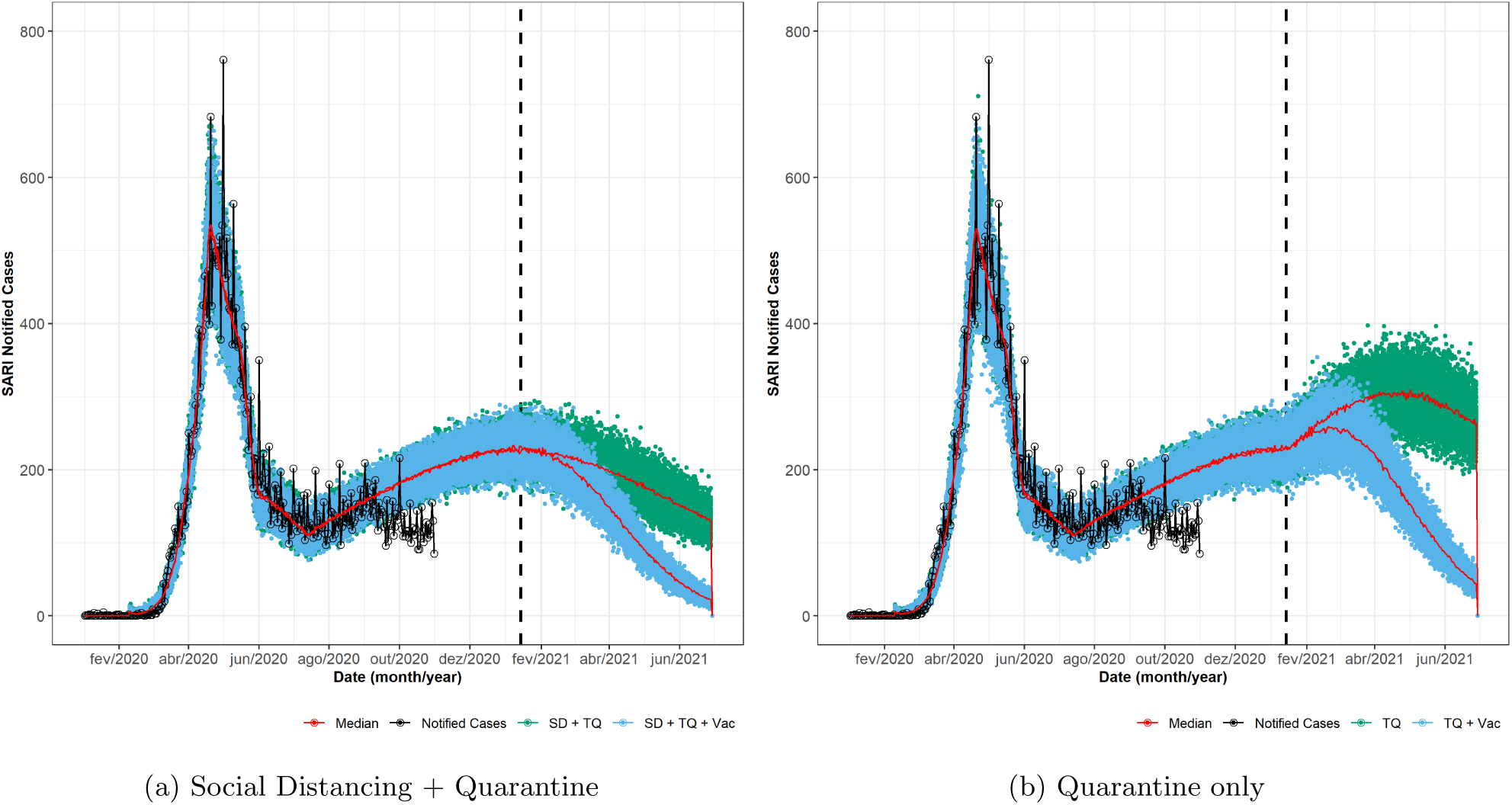
Comparison of the vaccination under different NPI. Black lines represent SARI notified cases. Green symbols represents a scenario without the vaccination, while the blue line represents the same conditions, but also applying the vaccine at 15 January 2020

## 4 Discussion

The main objective of NPI interventions is to mitigate the effect of the pandemic for a proper health care attention to mild and severe cases. As shown by Fig.2 independently from the nature of the intervention (social distancing or isolation of cases), as expected and seen in many studies [1, 11–13], delaying the epidemic peak is a consequence of the reduction in transmission intensity.

When comparing the different interventions, the combination of social distancing and isolation showed the best results, as demonstrated in Fig.2. When comparing the isolation interventions, there is a huge difference between the isolation of both symptomatic and asymptomatic cases, and isolating only the symptomatic cases, with the first one being the most successful application. This result highlights the importance of an enforced isolation measure, as the asymptomatic cases also impact in the transmission dynamics. The correct identification and consequently isolation of these cases pose a problem which has been discussed in the actual pandemic [14, 15], in some cases, following the correct procedure to identify and isolate these cases were responsible for ending the pandemic [16]. The isolation of only the severe cases did not do much to alter the behavior of the pandemic, demonstrating the importance of having a model in which mild and severe cases are studied separately, as they have marked differences in their epidemiology [17, 18] besides having some studies indicating some similarities [19, 20]. The only case in which the isolation of only the symptomatic cases was effective, was when it was applied altogether with the social distancing of all age groups. Therefore, it is imperative to recognize the importance of transmission by asymptomatic individuals.

Comparing the applied social distancing measures, results here show a very marked difference between the isolation of all age groups against the isolation of only young or elderly individuals and the severity of SARS-CoV-2 among elderly individuals higher than younger individuals [18, 20]. However, there must be a very careful distinction between the severity of cases and the epidemiological dynamic imposed by the different groups, the isolation of only the elder individuals is not sufficient to significantly halter the pandemic. As shown in our model, isolating the elderly group may give a false impression of protection to these individuals, as this intervention is not sufficient to effectively stop the epidemic, they are not effectively protected and still being subjected to be infected, and therefore, develop a severe case. Therefore, only the social distancing of all age groups at an early stage acts to avoid severe cases.

The social distancing of all age groups had similar performance compared to the isolation of both symptomatic and asymptomatic cases, as shown by Fig.2. This interesting result indicates that the early recognition and application of broad interventions to the population are the most effective measures to be studied. That is, acting with the objective of diminishing the number of possible infections through social distancing of healthy individuals or at the successful isolation of infected individuals is, as expected, equally important. The most important observation to be taken into account is that this intervention should be broad. In regard to the social distancing, all age groups should be taken into account, in agreement with other modeling studies [11, 12]. Regarding the isolation intervention, all cases should be included in the measure, including asymptomatic cases, which can only be reached through successful testing. This highlights the importance of mass testing individuals exposed to the SARS-CoV-2 pandemic.

Despite the great number of interventions, either a social distancing or isolation intervention, the best approach is clearly the combination of both measures. This is shown in 2 where the SD-A intervention combined with the TQ isolation measure produced the best results.

Despite all of the interventions, combined or not, act in a way to suppress the pandemic with different success rates, it is important to remember that there is a growing concern about the social and economic distress of a population during interventions [21, 22]. It is imperative to also observe and develop pharmaceutical interventions that are capable of reducing and really ending the posed a threat by the virus. Also, initiatives such as the vaccines being developed and the fundamental understanding of how the virus acts biologically are important to this end. Therefore, it is important to model beyond the dynamics of only non-pharmaceutical interventions.

Non-pharmaceutical interventions also demonstrate through Fig.4 that they have the merit of controlling the direction, evolution, and severity of the pandemic and should be studied and applied whenever possible. However, as shown by Figs.5 and 6, pharmaceutical and non-pharmaceutical interventions need to be considered altogether during the pandemic. Considering these results, it is clear that the vaccine has a long-term effect on the population. The vaccination schedule in this work is likely optimistic, mainly regarding the period of only 15 days between each phase and the vaccine requiring only one dose to be effective, while works in the literature already point out that two doses are going to be the required minimum for a good number of vaccine candidates [23, 24]. However, we wanted to test how a large, focused, and effective vaccination schedule would behave, and we see that not even a program with these characteristics is not subjected to the need of allowing NPI to be taken into account.

In all scenarios, the phased rollout of the vaccination program should be along with maintaining social distancing and case isolation. Abandoning the quarantine shows to be a most critical scenario, in which there is a considerable increase in the number of hospitalizations. The only condition where the pandemic maintains its downward strategy during the vaccination program is combining social distancing and quarantine. The scenario in which social distancing is more flexible shows an increase in the number of hospitalizations.

This is a crucial moment to study and show that we must yet consider the application of strict interventions of social distancing, isolation, and vaccination as the risk of SARS-CoV-2 transmission is present in multiple countries. The modelling in this work shows that effective control of the COVID-19 pandemic requires a combination of these efforts.

## Data Availability

All SARI and ARI notification data are publicly available at OpenDataSUS database, maintained by
the Ministry of Health, located at https://opendatasus.saude.gov.br/.

## Data Availability Statement

All SARI and ARI notification data are publicly available at OpenDataSUS database, maintained by the Ministry of Health, located at https://opendatasus.saude.gov.br/.

